## Supplementary material for "Protocol for the systematic review of the *Pneumocystis jirovecii*-associated pneumonia in non-HIV immunocompromised patients": S2 File. Sample search strategy for PubMed.

Supplementary file—appendix

S2 File. Sample search strategy for PubMed.

Available via <https://pubmed.ncbi.nlm.nih.gov>

Search date: 1 May 2023

Articles retrieved: 616.

| Search | Query | Results |
| --- | --- | --- |
| #1 | ((((“Pneumocystis jirovecii”) OR (“Pneumonia, Pneumocystis”) OR (“Pneumocystis Pneumonias”) OR (“Pneumonias, Pneumocystis”) OR (“Pneumonia, Pneumocystis jirovecii”) OR (“Pneumocystosis”) OR (“Pneumocystoses”) OR (“Pneumonia, Interstitial Plasma Cell”) OR (“Pneumocystis carinii Pneumonia”) OR (“Pneumocystis Pneumonia”)))) | [12,218](https://pubmed.ncbi.nlm.nih.gov/?term=%28%28%28%28%22Pneumocystis+jirovecii%22%29+OR+%28%22Pneumonia%2C+Pneumocystis%22%29+OR+%28%22Pneumocystis+Pneumonias%22%29+OR+%28%22Pneumonias%2C+Pneumocystis%22%29+OR+%28%22Pneumonia%2C+Pneumocystis+jirovecii%22%29+OR+%28%22Pneumocystosis%22%29+OR+%28%22Pneumocystoses%22%29+OR+%28%22Pneumonia%2C+Interstitial+Plasma+Cell%22%29+OR+%28%22Pneumocystis+carinii+Pneumonia%22%29+OR+%28%22Pneumocystis+Pneumonia%22%29%29%29%29&ac=no&sort=relevance) |
| #2 | (((("Pneumocystis jirovecii"[Title/Abstract]) OR ("Pneumonia, Pneumocystis"[Title/Abstract]) OR ("Pneumocystis Pneumonias"[Title/Abstract]) OR ("Pneumonias, Pneumocystis"[Title/Abstract]) OR ("Pneumonia, Pneumocystis jirovecii"[Title/Abstract]) OR ("Pneumocystosis"[Title/Abstract]) OR ("Pneumocystoses"[Title/Abstract]) OR ("Pneumonia, Interstitial Plasma Cell"[Title/Abstract]) OR ("Pneumocystis carinii Pneumonia"[Title/Abstract]) OR ("Pneumocystis Pneumonia"[Title/Abstract])))) | [8,361](https://pubmed.ncbi.nlm.nih.gov/?term=%28%28%28%28%22Pneumocystis+jirovecii%22%5BTitle%2FAbstract%5D%29+OR+%28%22Pneumonia%2C+Pneumocystis%22%5BTitle%2FAbstract%5D%29+OR+%28%22Pneumocystis+Pneumonias%22%5BTitle%2FAbstract%5D%29+OR+%28%22Pneumonias%2C+Pneumocystis%22%5BTitle%2FAbstract%5D%29+OR+%28%22Pneumonia%2C+Pneumocystis+jirovecii%22%5BTitle%2FAbstract%5D%29+OR+%28%22Pneumocystosis%22%5BTitle%2FAbstract%5D%29+OR+%28%22Pneumocystoses%22%5BTitle%2FAbstract%5D%29+OR+%28%22Pneumonia%2C+Interstitial+Plasma+Cell%22%5BTitle%2FAbstract%5D%29+OR+%28%22Pneumocystis+carinii+Pneumonia%22%5BTitle%2FAbstract%5D%29+OR+%28%22Pneumocystis+Pneumonia%22%5BTitle%2FAbstract%5D%29%29%29%29&ac=no&sort=relevance) |
| #3 | (((("Immunocompromised Host") OR ("Host, Immunocompromised") OR ("Hosts, Immunocompromised") OR ("Immunocompromised Hosts") OR ("Immunocompromised Patient") OR ("Immunocompromised Patients") OR ("Patient, Immunocompromised") OR ("Patients, Immunocompromised") OR ("Immunosuppressed Host") OR ("Host, Immunosuppressed") OR ("Hosts, Immunosuppressed") OR ("Immunosuppressed Hosts")))) | [42,567](https://pubmed.ncbi.nlm.nih.gov/?term=%28%28%28%28%22Immunocompromised+Host%22%29+OR+%28%22Host%2C+Immunocompromised%22%29+OR+%28%22Hosts%2C+Immunocompromised%22%29+OR+%28%22Immunocompromised+Hosts%22%29+OR+%28%22Immunocompromised+Patient%22%29+OR+%28%22Immunocompromised+Patients%22%29+OR+%28%22Patient%2C+Immunocompromised%22%29+OR+%28%22Patients%2C+Immunocompromised%22%29+OR+%28%22Immunosuppressed+Host%22%29+OR+%28%22Host%2C+Immunosuppressed%22%29+OR+%28%22Hosts%2C+Immunosuppressed%22%29+OR+%28%22Immunosuppressed+Hosts%22%29%29%29%29&ac=no&sort=relevance) |
| #4 | (((("Immunocompromised Host"[Title/Abstract]) OR ("Host, Immunocompromised"[Title/Abstract]) OR ("Hosts, Immunocompromised"[Title/Abstract]) OR ("Immunocompromised Hosts"[Title/Abstract]) OR ("Immunocompromised Patient"[Title/Abstract]) OR ("Immunocompromised Patients"[Title/Abstract]) OR ("Patient, Immunocompromised"[Title/Abstract]) OR ("Patients, Immunocompromised"[Title/Abstract]) OR ("Immunosuppressed Host"[Title/Abstract]) OR ("Host, Immunosuppressed"[Title/Abstract]) OR ("Hosts, Immunosuppressed"[Title/Abstract]) OR ("Immunosuppressed Hosts"[Title/Abstract])))) | [25,008](https://pubmed.ncbi.nlm.nih.gov/?term=%28%28%28%28%22Immunocompromised+Host%22%5BTitle%2FAbstract%5D%29+OR+%28%22Host%2C+Immunocompromised%22%5BTitle%2FAbstract%5D%29+OR+%28%22Hosts%2C+Immunocompromised%22%5BTitle%2FAbstract%5D%29+OR+%28%22Immunocompromised+Hosts%22%5BTitle%2FAbstract%5D%29+OR+%28%22Immunocompromised+Patient%22%5BTitle%2FAbstract%5D%29+OR+%28%22Immunocompromised+Patients%22%5BTitle%2FAbstract%5D%29+OR+%28%22Patient%2C+Immunocompromised%22%5BTitle%2FAbstract%5D%29+OR+%28%22Patients%2C+Immunocompromised%22%5BTitle%2FAbstract%5D%29+OR+%28%22Immunosuppressed+Host%22%5BTitle%2FAbstract%5D%29+OR+%28%22Host%2C+Immunosuppressed%22%5BTitle%2FAbstract%5D%29+OR+%28%22Hosts%2C+Immunosuppressed%22%5BTitle%2FAbstract%5D%29+OR+%28%22Immunosuppressed+Hosts%22%5BTitle%2FAbstract%5D%29%29%29%29&ac=no&sort=relevance) |
| #5 | ((("Pneumonia") OR ("Pneumonias") OR ("Lobar Pneumonia") OR ("Lobar Pneumonias") OR ("Pneumonias, Lobar") OR ("Pneumonia, Lobar") OR ("Experimental Lung Inflammation") OR ("Experimental Lung Inflammations") OR ("Inflammation, Experimental Lung") OR ("Lung Inflammation, Experimental") OR ("Lung Inflammations, Experimental") OR ("Pneumonitis") OR ("Pneumonitides") OR ("Pulmonary Inflammation") OR ("Inflammation, Pulmonary") OR ("Inflammations, Pulmonary") OR ("Pulmonary Inflammations") OR ("Lung Inflammation") OR ("Inflammation, Lung") OR ("Inflammations, Lung") OR ("Lung Inflammations"))) | [459,577](https://pubmed.ncbi.nlm.nih.gov/?term=%28%28%28%22Pneumonia%22%29+OR+%28%22Pneumonias%22%29+OR+%28%22Lobar+Pneumonia%22%29+OR+%28%22Lobar+Pneumonias%22%29+OR+%28%22Pneumonias%2C+Lobar%22%29+OR+%28%22Pneumonia%2C+Lobar%22%29+OR+%28%22Experimental+Lung+Inflammation%22%29+OR+%28%22Experimental+Lung+Inflammations%22%29+OR+%28%22Inflammation%2C+Experimental+Lung%22%29+OR+%28%22Lung+Inflammation%2C+Experimental%22%29+OR+%28%22Lung+Inflammations%2C+Experimental%22%29+OR+%28%22Pneumonitis%22%29+OR+%28%22Pneumonitides%22%29+OR+%28%22Pulmonary+Inflammation%22%29+OR+%28%22Inflammation%2C+Pulmonary%22%29+OR+%28%22Inflammations%2C+Pulmonary%22%29+OR+%28%22Pulmonary+Inflammations%22%29+OR+%28%22Lung+Inflammation%22%29+OR+%28%22Inflammation%2C+Lung%22%29+OR+%28%22Inflammations%2C+Lung%22%29+OR+%28%22Lung+Inflammations%22%29%29%29&ac=no&sort=relevance) |
| #6 | ((("Pneumonia"[Title/Abstract]) OR ("Pneumonias"[Title/Abstract]) OR ("Lobar Pneumonia"[Title/Abstract]) OR ("Lobar Pneumonias"[Title/Abstract]) OR ("Pneumonias, Lobar"[Title/Abstract]) OR ("Pneumonia, Lobar"[Title/Abstract]) OR ("Experimental Lung Inflammation"[Title/Abstract]) OR ("Experimental Lung Inflammations"[Title/Abstract]) OR ("Inflammation, Experimental Lung"[Title/Abstract]) OR ("Lung Inflammation, Experimental"[Title/Abstract]) OR ("Lung Inflammations, Experimental"[Title/Abstract]) OR ("Pneumonitis"[Title/Abstract]) OR ("Pneumonitides"[Title/Abstract]) OR ("Pulmonary Inflammation"[Title/Abstract]) OR ("Inflammation, Pulmonary"[Title/Abstract]) OR ("Inflammations, Pulmonary"[Title/Abstract]) OR ("Pulmonary Inflammations"[Title/Abstract]) OR ("Lung Inflammation"[Title/Abstract]) OR ("Inflammation, Lung"[Title/Abstract]) OR ("Inflammations, Lung"[Title/Abstract]) OR ("Lung Inflammations"[Title/Abstract]))) | [185,749](https://pubmed.ncbi.nlm.nih.gov/?term=%28%28%28%22Pneumonia%22%5BTitle%2FAbstract%5D%29+OR+%28%22Pneumonias%22%5BTitle%2FAbstract%5D%29+OR+%28%22Lobar+Pneumonia%22%5BTitle%2FAbstract%5D%29+OR+%28%22Lobar+Pneumonias%22%5BTitle%2FAbstract%5D%29+OR+%28%22Pneumonias%2C+Lobar%22%5BTitle%2FAbstract%5D%29+OR+%28%22Pneumonia%2C+Lobar%22%5BTitle%2FAbstract%5D%29+OR+%28%22Experimental+Lung+Inflammation%22%5BTitle%2FAbstract%5D%29+OR+%28%22Experimental+Lung+Inflammations%22%5BTitle%2FAbstract%5D%29+OR+%28%22Inflammation%2C+Experimental+Lung%22%5BTitle%2FAbstract%5D%29+OR+%28%22Lung+Inflammation%2C+Experimental%22%5BTitle%2FAbstract%5D%29+OR+%28%22Lung+Inflammations%2C+Experimental%22%5BTitle%2FAbstract%5D%29+OR+%28%22Pneumonitis%22%5BTitle%2FAbstract%5D%29+OR+%28%22Pneumonitides%22%5BTitle%2FAbstract%5D%29+OR+%28%22Pulmonary+Inflammation%22%5BTitle%2FAbstract%5D%29+OR+%28%22Inflammation%2C+Pulmonary%22%5BTitle%2FAbstract%5D%29+OR+%28%22Inflammations%2C+Pulmonary%22%5BTitle%2FAbstract%5D%29+OR+%28%22Pulmonary+Inflammations%22%5BTitle%2FAbstract%5D%29+OR+%28%22Lung+Inflammation%22%5BTitle%2FAbstract%5D%29+OR+%28%22Inflammation%2C+Lung%22%5BTitle%2FAbstract%5D%29+OR+%28%22Inflammations%2C+Lung%22%5BTitle%2FAbstract%5D%29+OR+%28%22Lung+Inflammations%22%5BTitle%2FAbstract%5D%29%29%29&ac=no&sort=relevance) |
| #7 | (((((("Pneumocystis jirovecii"[Title/Abstract]) OR ("Pneumonia, Pneumocystis"[Title/Abstract]) OR ("Pneumocystis Pneumonias"[Title/Abstract]) OR ("Pneumonias, Pneumocystis"[Title/Abstract]) OR ("Pneumonia, Pneumocystis jirovecii"[Title/Abstract]) OR ("Pneumocystosis"[Title/Abstract]) OR ("Pneumocystoses"[Title/Abstract]) OR ("Pneumonia, Interstitial Plasma Cell"[Title/Abstract]) OR ("Pneumocystis carinii Pneumonia"[Title/Abstract]) OR ("Pneumocystis Pneumonia"[Title/Abstract]))))) AND ((((("Immunocompromised Host"[Title/Abstract]) OR ("Host, Immunocompromised"[Title/Abstract]) OR ("Hosts, Immunocompromised"[Title/Abstract]) OR ("Immunocompromised Hosts"[Title/Abstract]) OR ("Immunocompromised Patient"[Title/Abstract]) OR ("Immunocompromised Patients"[Title/Abstract]) OR ("Patient, Immunocompromised"[Title/Abstract]) OR ("Patients, Immunocompromised"[Title/Abstract]) OR ("Immunosuppressed Host"[Title/Abstract]) OR ("Host, Immunosuppressed"[Title/Abstract]) OR ("Hosts, Immunosuppressed"[Title/Abstract]) OR ("Immunosuppressed Hosts"[Title/Abstract])))))) AND (((("Pneumonia") OR ("Pneumonias") OR ("Lobar Pneumonia") OR ("Lobar Pneumonias") OR ("Pneumonias, Lobar") OR ("Pneumonia, Lobar") OR ("Experimental Lung Inflammation") OR ("Experimental Lung Inflammations") OR ("Inflammation, Experimental Lung") OR ("Lung Inflammation, Experimental") OR ("Lung Inflammations, Experimental") OR ("Pneumonitis") OR ("Pneumonitides") OR ("Pulmonary Inflammation") OR ("Inflammation, Pulmonary") OR ("Inflammations, Pulmonary") OR ("Pulmonary Inflammations") OR ("Lung Inflammation") OR ("Inflammation, Lung") OR ("Inflammations, Lung") OR ("Lung Inflammations")))) | [616](https://pubmed.ncbi.nlm.nih.gov/?term=%28%28%28%28%28%28%22Pneumocystis+jirovecii%22%5BTitle%2FAbstract%5D%29+OR+%28%22Pneumonia%2C+Pneumocystis%22%5BTitle%2FAbstract%5D%29+OR+%28%22Pneumocystis+Pneumonias%22%5BTitle%2FAbstract%5D%29+OR+%28%22Pneumonias%2C+Pneumocystis%22%5BTitle%2FAbstract%5D%29+OR+%28%22Pneumonia%2C+Pneumocystis+jirovecii%22%5BTitle%2FAbstract%5D%29+OR+%28%22Pneumocystosis%22%5BTitle%2FAbstract%5D%29+OR+%28%22Pneumocystoses%22%5BTitle%2FAbstract%5D%29+OR+%28%22Pneumonia%2C+Interstitial+Plasma+Cell%22%5BTitle%2FAbstract%5D%29+OR+%28%22Pneumocystis+carinii+Pneumonia%22%5BTitle%2FAbstract%5D%29+OR+%28%22Pneumocystis+Pneumonia%22%5BTitle%2FAbstract%5D%29%29%29%29%29+AND+%28%28%28%28%28%22Immunocompromised+Host%22%5BTitle%2FAbstract%5D%29+OR+%28%22Host%2C+Immunocompromised%22%5BTitle%2FAbstract%5D%29+OR+%28%22Hosts%2C+Immunocompromised%22%5BTitle%2FAbstract%5D%29+OR+%28%22Immunocompromised+Hosts%22%5BTitle%2FAbstract%5D%29+OR+%28%22Immunocompromised+Patient%22%5BTitle%2FAbstract%5D%29+OR+%28%22Immunocompromised+Patients%22%5BTitle%2FAbstract%5D%29+OR+%28%22Patient%2C+Immunocompromised%22%5BTitle%2FAbstract%5D%29+OR+%28%22Patients%2C+Immunocompromised%22%5BTitle%2FAbstract%5D%29+OR+%28%22Immunosuppressed+Host%22%5BTitle%2FAbstract%5D%29+OR+%28%22Host%2C+Immunosuppressed%22%5BTitle%2FAbstract%5D%29+OR+%28%22Hosts%2C+Immunosuppressed%22%5BTitle%2FAbstract%5D%29+OR+%28%22Immunosuppressed+Hosts%22%5BTitle%2FAbstract%5D%29%29%29%29%29%29+AND+%28%28%28%28%22Pneumonia%22%29+OR+%28%22Pneumonias%22%29+OR+%28%22Lobar+Pneumonia%22%29+OR+%28%22Lobar+Pneumonias%22%29+OR+%28%22Pneumonias%2C+Lobar%22%29+OR+%28%22Pneumonia%2C+Lobar%22%29+OR+%28%22Experimental+Lung+Inflammation%22%29+OR+%28%22Experimental+Lung+Inflammations%22%29+OR+%28%22Inflammation%2C+Experimental+Lung%22%29+OR+%28%22Lung+Inflammation%2C+Experimental%22%29+OR+%28%22Lung+Inflammations%2C+Experimental%22%29+OR+%28%22Pneumonitis%22%29+OR+%28%22Pneumonitides%22%29+OR+%28%22Pulmonary+Inflammation%22%29+OR+%28%22Inflammation%2C+Pulmonary%22%29+OR+%28%22Inflammations%2C+Pulmonary%22%29+OR+) |
