## Supplementary material for "Protocol for the systematic review of the *Pneumocystis jirovecii*-associated pneumonia in non-HIV immunocompromised patients": S3 File. PRISMA 2009 flow diagram

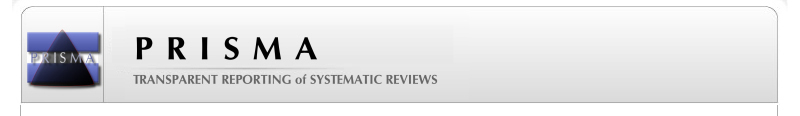
**PRISMA 2009 Flow Diagram**

**Screening**

**Included**

**Eligibility**

**Identification**

Records identified through database searching
(n =	)

Additional records identified through other sources
(n =	)

Records after duplicates removed
(n =	)

Records screened
(n =	)

Records excluded
(n =	)

Full-text articles assessed for eligibility
(n =	)

Full-text articles excluded, with reasons
(n =	)

Studies included in qualitative synthesis
(n =	)

Studies included in quantitative synthesis (meta-analysis)
(n =	)
