## Supplementary material for "Protocol for the systematic review of the *Pneumocystis jirovecii*-associated pneumonia in non-HIV immunocompromised patients": S4 File. Data extraction form for systematic review.

| Title: |  |
| --- | --- |
| Bibliographic Information: |  |
| **DATA COLLECTION ITEM** | **ITEM DESCRIPTION / SUB-GROUPS** |
| AUTHOR | Name(s) of the author(s) |
| TITLE | Title of the research paper |
| YEAR OF PUBLICATION | The year when the study was published |
| COUNTRY | The country where the study was conducted |

S4 Table: Data Extraction Form for PJP Systematic Review

| Methods: |  |
| --- | --- |
| **DATA COLLECTION ITEM** | **ITEM DESCRIPTION / SUB-GROUPS** |
| TYPE OF STUDY | Type of research design (e.g., randomized controlled trial, cohort study) |
| TIME PERIOD AND SETTING OF THE STUDY | Duration and location where the study took place |
| INTERVENTION | Type of treatment or prophylaxis for PJP |
| DESCRIPTION OF INTERVENTION | Detailed description of the PJP intervention |
| THE SETTING OF THE INTERVENTION | Healthcare setting or community-based setting |
| AIM/OBJECTIVES | Objectives related to PJP outcomes |
| PARTICIPANT CHARACTERISTICS | - Participants: Characteristics of individuals at risk for or diagnosed with PJP |
|  | - Comparator details: Description of the comparison group, if applicable |
|  | - Inclusion Criteria: Criteria for including participants in PJP studies |
|  | - Exclusion Criteria: Criteria for excluding participants from PJP studies |
| OUTCOME MEASURES | PJP-specific outcomes were measured in the study. |
| SOURCE OF DATA | Information about where the data for PJP outcomes was sourced. |
| DATA ANALYSIS | Methods used for analyzing PJP-related data. |

| Results: |  |
| --- | --- |
| **DATA COLLECTION ITEM** | **ITEM DESCRIPTION / SUB-GROUPS** |
| STUDY PARAMETERS | Key parameters specific to PJP |
| OUTCOMES | Mean values for PJP-related outcomes |
|  | Mean differences between comparator groups in the context of PJP studies |
| CHARACTERIZING HETEROGENEITY | Assessment of heterogeneity in PJP study results |

| Discussion: |  |
| --- | --- |
| **DATA COLLECTION ITEM** | **ITEM DESCRIPTION / SUB-GROUPS** |
| STUDY FINDINGS | Summary of key PJP study findings |
| LIMITATIONS | Limitations of the study, especially regarding PJP research |
| GENERALISABILITY | Generalizability of the study findings to broader PJP populations or settings |
| CURRENT KNOWLEDGE | Alignment of PJP findings with existing knowledge |
| CHANGES IN DIAGNOSTIC TESTS | Discussion of any changes or implications for PJP diagnostic tests based on the study |
| CONCLUSION | One-sentence conclusion of the study. |
