## Supplementary material for "Protocol for the systematic review of the *Pneumocystis jirovecii*-associated pneumonia in non-HIV immunocompromised patients": S5 File. PROSPERO registration.

To enable PROSPERO to focus on COVID-19 submissions, this registration record has undergone basic automated checks for eligibility and is published exactly as submitted. PROSPERO has never provided peer review, and usual checking by the PROSPERO team does not endorse content. Therefore, automatically published records should be treated as any other PROSPERO registration. Further detail is provided here.

### Citation

Mauricio Ernesto Orozco Ugarriza, Yuranis Eugenia Rodger Cervantes, Yenifer Olivo Martínez. Pneumocystis jirovecii-associated pneumonia in non-HIV immunocompromised patients. PROSPERO 2023 CRD42023429112 Available from: https://www.crd.york.ac.uk/prospero/display_record.php?ID=CRD42023429112

### Review question

P: What is the incidence, clinical characteristics, risk factors, and therapeutic management of Pneumocystis jirovecii-associated pneumonia in non-HIV immunocompromised patients?

I: Incidence, clinical characteristics, risk factors, and therapeutic management.

E: No specific intervention is applied.

C: No direct comparison is applicable.

O: To determine the incidence, describe the clinical characteristics, identify risk factors, and evaluate therapeutic management options for Pneumocystis jirovecii-associated pneumonia in non-HIV immunocompromised patients.

### Searches

The search strategy will be conducted comprehensively across electronic databases, including PubMed, Google Scholar, and ScienceDirect. Controlled terms (also known as Medical Subject Headings or MeSH terms) and relevant keywords will be used to identify relevant studies related to Pneumocystis jirovecii-associated pneumonia in immunocompromised patients without a diagnosis of HIV.

The search strategy will be constructed using combinations of the following terms:

Pneumonia

Pneumocystis jirovecii

Immunocompromised patients

Without HIV diagnosis

Incidence

Clinical characteristics

Risk factors

Therapeutic management

Boolean operators (such as AND, OR) and truncations will be used, as per the specificities of each database, to expand and refine the search. The search will be limited to studies published between 2010 and 2022, aiming to obtain the most up-to-date evidence on the topic of interest. Only studies published in English will be included. However, efforts will be made to identify and access relevant studies published in other languages, if possible.The details of the search strategy, including the terms used and filters applied, will be recorded to ensure transparency and reproducibility of the systematic review.

### Types of study to be included

The types of studies eligible for inclusion in this systematic review are original articles that declare an epidemiological research design of an analytical observational nature, such as cohort studies, case-control studies, cross-sectional studies, and retrospective studies. These studies should investigate Pneumocystis jirovecii-associated pneumonia in immunocompromised patients without a diagnosis of HIV. The inclusion criteria include the study design (analytical observational studies), study population (immunocompromised patients without HIV), and outcomes of interest (incidence, clinical characteristics, risk factors, and therapeutic management). Excluded studies are those that do not specifically address the topic, use different study designs, or are not available in English or accessible languages. There are no additional restrictions on eligible study types for this review.

### Condition or domain being studied

The systematic review focuses on Pneumocystis jirovecii-associated pneumonia in immunocompromised patients without a diagnosis of HIV. Pneumocystis jirovecii-associated pneumonia is a lung infection caused by the fungus Pneumocystis jirovecii. It primarily affects individuals with weakened immune systems, such as those who have undergone organ transplantation, are receiving immunosuppressive treatment, or have autoimmune diseases.

This condition is of particular importance due to its impact on the morbidity and mortality of immunocompromised patients. Symptoms of Pneumocystis jirovecii-associated pneumonia include difficulty breathing, dry cough, fever, and fatigue. Without proper diagnosis and treatment, the disease can progress rapidly and be potentially life-threatening.

The systematic review aims to analyze the incidence of Pneumocystis jirovecii-associated pneumonia in immunocompromised patients without a diagnosis of HIV, as well as describe the clinical characteristics of the disease, identify associated risk factors, and evaluate available therapeutic management options. The goal is to gather existing evidence to better understand this condition and provide relevant information for clinical decision-making and the development of effective prevention and treatment strategies.

### Participants/population

The participants or populations being studied in this systematic review are immunocompromised patients without a diagnosis of HIV who have been diagnosed with Pneumocystis jirovecii-associated pneumonia. The inclusion criteria for participants include individuals of any age or gender who meet the defined criteria for immunocompromised status and have been diagnosed with Pneumocystis jirovecii-associated pneumonia. Patients with a confirmed diagnosis of HIV will be excluded from this review. There are no specific restrictions based on geographic location or any other demographic factors. Studies that include participants who do not meet the defined criteria or focus solely on HIV-infected individuals will be excluded.

### Intervention(s), exposure(s)

The interventions or exposures under review in this SR pertain to P. jirovecii-associated pneumonia in immunocompromised individuals without HIV. The review aims to examine the incidence, clinical characteristics, risk factors, and therapeutic management of this condition. The specific interventions or exposures of interest are as follows:

Incidence: Will assess the occurrence rate of P. jirovecii-associated pneumonia in immunocompromised individuals without HIV, focusing on the identification of new cases within a defined population or time period.

Clinical Characteristics: Will investigate the signs, symptoms, diagnostic criteria, and disease progression patterns associated with P. jirovecii-associated pneumonia in the target population. This includes descriptions of radiological findings, laboratory and clinical presentations.

Risk Factors: The review aims to identify the factors that contribute to the development or increased risk of P. jirovecii-associated pneumonia in immunocompromised individuals without HIV. This may encompass underlying medical conditions, immunosuppressive therapies, environmental exposures, or genetic predisposition.

Therapeutic Management: Will analyze various approaches to the treatment and management of pneumonia in the target population. This includes pharmacological interventions, supportive care measures, preventive and clinical management strategies.

The inclusion criteria will focus on studies that provide comprehensive descriptions of these interventions or exposures, enabling reproducibility and assessment of their applicability in other settings.

### Comparator(s)/control

Not applicable

### Context

The inclusion criteria for participant selection in this systematic review are as follows:

Study Participants: The review will focus on immunocompromised individuals without a diagnosis of HIV. This includes patients with conditions such as organ transplantation, hematological malignancies, solid tumors, autoimmune disorders, chronic use of immunosuppressive medications, or any other underlying condition that leads to immunosuppression.

Study Setting: The review will consider studies conducted in various healthcare settings, including hospitals, clinics, and community healthcare facilities. There are no restrictions based on the setting of the study. However, studies conducted in specific settings that are relevant to the research question, such as intensive care units or specialized immunology clinics, will be given particular attention.

Study Design: The review will include original research articles reporting on observational epidemiological study designs, such as cohort studies, case-control studies, cross-sectional studies, and retrospective studies. These study designs are well-suited to assess the incidence, characteristics, risk factors, and therapeutic management of Pneumocystis jirovecii-associated pneumonia in immunocompromised individuals without HIV.

Regarding other relevant characteristics, studies conducted exclusively in hospital accident and emergency departments will not be included. Similarly, research limited to low- and middle-income countries will not be a restriction for inclusion in this review. Instead, studies conducted in various geographical locations and settings will be considered to provide a comprehensive understanding of the topic.

### Main outcome(s)

The primary outcomes of this systematic review include the following pre-specified and important results:

Incidence of Pneumocystis jirovecii-associated pneumonia: The review aims to determine the occurrence rate of Pneumocystis jirovecii-associated pneumonia in immunocompromised individuals without a diagnosis of HIV.

Clinical characteristics of Pneumocystis jirovecii-associated pneumonia: The review seeks to describe the clinical features associated with Pneumocystis jirovecii-associated pneumonia in immunocompromised patients.

Risk factors for Pneumocystis jirovecii-associated pneumonia: The review aims to identify factors that increase the likelihood of developing Pneumocystis jirovecii-associated pneumonia in immunocompromised individuals without HIV.

Therapeutic management of Pneumocystis jirovecii-associated pneumonia: The review seeks to evaluate treatment approaches used for Pneumocystis jirovecii-associated pneumonia in immunocompromised patients.

These outcomes will be measured using data reported in the included studies, including documented cases, clinical assessments, laboratory tests, imaging findings, treatment regimens, and patient outcomes. The time frame for outcome measurement will vary depending on the information provided in the included studies.

#### Measures of effect

The effect measures for the main outcomes of this systematic review, focusing on Pneumocystis jirovecii-associated pneumonia in immunocompromised individuals without HIV, will vary based on the data reported in the included studies. The main outcomes include incidence, clinical characteristics, risk factors, and therapeutic management.

For the incidence of Pneumocystis jirovecii-associated pneumonia, the effect measure will be the cumulative incidence rate or the number of new cases per person-time at risk, providing an estimate of the risk of developing the condition.

Regarding clinical characteristics, risk factors, and therapeutic management, the effect measures will primarily consist of odds ratios (OR) or risk ratios (RR) depending on the study design. These measures quantify the association between the exposure or intervention of interest and the examined outcomes.

Additional effect measures such as risk difference (RD) or number needed to treat (NNT) may be used if relevant data are available in the included studies, providing insights into absolute differences in outcomes between different groups or interventions.

The selection of specific effect measures will depend on the extracted data and the statistical methods employed in the analysis. The chosen measures will ensure appropriate and meaningful comparisons across studies, considering the study designs and available data.

### Additional outcome(s)

The pre-specified additional secondary outcomes of this systematic review will include:

Mortality related to Pneumocystis jirovecii-associated pneumonia in immunocompromised patients without a diagnosis of HIV.

Respiratory complications associated with pneumonia, such as the need for mechanical ventilation, acute respiratory failure, or the development of acute respiratory distress syndrome.

Length of hospital stay related to pneumonia.

Recurrence of Pneumocystis jirovecii-associated pneumonia.

Adverse effects associated with the therapeutic management of pneumonia, such as allergic reactions to antiparasitic medications.

A detailed assessment of these secondary outcomes will be conducted, including how they are defined and measured, as well as the timing of relevant measurements. These outcomes will provide additional information on the severity of the disease, the effectiveness of therapeutic interventions, and the impact on patients' health.

In case there are no pre-specified additional secondary outcomes, it will be stated as "Not applicable" in relation to this systematic review.

#### Measures of effect

The effect measures for the additional outcomes of this systematic review will be specified as follows:

Mortality related to Pneumocystis jirovecii-associated pneumonia in immunocompromised patients without a diagnosis of HIV: The effect measure will be reported as relative risks (RR) or odds ratios (OR) with corresponding confidence intervals (CI).

Respiratory complications associated with pneumonia: The effect measure will be reported as relative risks (RR) or odds ratios (OR) with corresponding confidence intervals (CI).

Length of hospital stay related to pneumonia: The effect measure will be reported as mean differences (MD) or standardized mean differences (SMD) with corresponding confidence intervals (CI).

Recurrence of Pneumocystis jirovecii-associated pneumonia: The effect measure will be reported as risk difference (RD) or hazard ratio (HR) with corresponding confidence intervals (CI).

Adverse effects associated with the therapeutic management of pneumonia: The effect measure will be reported as relative risks (RR) or odds ratios (OR) with corresponding confidence intervals (CI).

These effect measures will allow for the evaluation of the impact of Pneumocystis jirovecii-associated pneumonia on various outcomes and the effectiveness of different therapeutic interventions.

### Data extraction (selection and coding)

In the systematic review, the data extraction process will involve the following steps:

Study selection: Multiple reviewers will independently apply the eligibility criteria and select studies for inclusion in the systematic review. Disagreements between reviewers' judgments will be resolved through discussion and consensus. Study inclusion/exclusion decisions will be recorded using a suitable tool or format.

Data extraction: Relevant data will be extracted from study documents, including information on the first author and year of publication, geographical region of the study, study design, methodology, participant demographics, baseline characteristics, outcome measures, and relevant laboratory results.

Multiple individuals will be involved in data extraction or checking to ensure accuracy and minimize errors.

Disagreements between data extractors will be resolved through discussion and consensus.

Efforts will be made to contact study investigators to request unreported data or additional details for missing data.

The extracted data will be recorded using a suitable tool or format, such as an Excel spreadsheet.

The systematic review team will adhere to established guidelines and good practices to ensure a rigorous and transparent data extraction process. This will involve independent assessment, resolution of disagreements, and thorough documentation of study selection and data extraction procedures.

### Risk of bias (quality) assessment

For the systematic review, the method of assessing risk of bias or study quality will involve the following steps:

Independent assessment: Two reviewers will independently evaluate the methodological quality of the included studies or literature.

Disagreement resolution: Any disagreements between reviewers' assessments will be resolved through discussion and consensus. In cases where consensus cannot be reached, a third reviewer will be involved to reach a final agreement.

Study selection and data extraction: Two reviewers will independently assess the search results and select high-quality literature for inclusion in the review. Multiple reviewers will be involved in the data extraction process to minimize the risk of errors.

Blinding: The study selection, data extraction, and risk of bias assessment will be performed without blinding of the assessors to study authors or the journal of publication.

These measures are implemented to ensure the rigor and reliability of the review process. By involving multiple independent reviewers and resolving disagreements through discussion and consensus, the risk of bias in the included studies will be assessed in a transparent and comprehensive manner.

### Strategy for data synthesis

We planned to synthesize data using a narrative synthesis approach, which will involve summarizing the interventions, outcomes of interest, population characteristics, and study quality. If possible, descriptive statistics will be employed to analyze the results. A narrative synthesis of findings, including the main findings and limitations, will be prepared. If appropriate, we may group these findings into categories.

This systematic review will be performed in accordance with the guidance specified in the PRISMA statement for reporting systematic reviews of health care interventions. The PRISMA guidelines ensure transparency and rigor in the review process.

The narrative synthesis will provide a comprehensive overview of the available evidence. It will involve synthesizing and summarizing the data extracted from the included studies, focusing on the clinical characteristics, risk factors, and therapeutic management of Pneumocystis jirovecii-associated pneumonia in immunocompromised individuals without HIV. The narrative synthesis will also consider the methodological quality of the studies and any limitations identified.

Overall, the narrative synthesis will allow for a comprehensive understanding of the topic by presenting a cohesive narrative that integrates the findings from the included studies.

### Analysis of subgroups or subsets

For the systematic review on "Pneumocystis jirovecii-associated pneumonia in immunocompromised individuals without HIV," we do not have any planned investigation of subgroups or subsets. The aim of this review is to provide a comprehensive synthesis of the available evidence regarding the clinical characteristics, risk factors, and therapeutic management of this condition in the specified population. Since no specific differences or effect modifiers are anticipated in the included studies, there is no need for subgroup analysis.

The focus of this review is to analyze and synthesize the overall evidence from the included studies. We will not be conducting separate analyses based on different types of studies or individual characteristics. Instead, we will consider the collective findings and characteristics of the included studies as a whole.

The planned analytic approach for this systematic review involves a narrative synthesis, as previously described. We will summarize and present the findings, interventions, outcomes, and population characteristics of the included studies in a comprehensive manner. We will not be employing meta-regression, tests of interaction, or statistical modeling since the objective is to provide a qualitative synthesis rather than a quantitative analysis.

The primary goal is to present a comprehensive overview of the topic, identify key findings, and discuss their implications.

### Contact details for further information

Mauricio Ernesto Orozco Ugarriza

### Organisational affiliation of the review

Universidad de San Buenaventura Cartagena

https://usbcartagena.edu.co/

### Review team members and their organisational affiliations

Professor Mauricio Ernesto Orozco Ugarriza. Universidad de San Buenaventura Cartagena, Colombia

Ms Yuranis Eugenia Rodger Cervantes. Universidad de San Buenaventura Cartagena, Colombia

Professor Yenifer Olivo Martínez. Universidad de San Buenaventura Cartagena, Colombia

### Type and method of review

Narrative synthesis, Systematic review

### Anticipated or actual start date

01 September 2017

### Anticipated completion date

30 June 2023

### Funding sources/sponsors

None

### Conflicts of interest

### Language

English

### Country

Colombia

### Stage of review

Review Ongoing

### Subject index terms status

Subject indexing assigned by CRD

### Subject index terms

Humans; Immunocompromised Host; Incidence; Pneumocystis carinii; Pneumonia, Pneumocystis; Risk Factors

### Date of registration in PROSPERO

05 June 2023

### Date of first submission

25 May 2023

### Stage of review at time of this submission

| Stage | Started | Completed |
|---|---|---|
| Preliminary searches | Yes | No |
| Piloting of the study selection process | Yes | No |
| Formal screening of search results against eligibility criteria | Yes | No |
| Data extraction | No | No |
| Risk of bias (quality) assessment | No | No |
| Data analysis | No | No |

*The record owner confirms that the information they have supplied for this submission is accurate and complete and they understand that deliberate provision of inaccurate information or omission of data may be construed as scientific misconduct.*

*The record owner confirms that they will update the status of the review when it is completed and will add publication details in due course.*

### Versions

05 June 2023

05 June 2023
